# Features of creatine-kinase in COVID-19 patients with different ages, clinical types and outcomes: A cohort study

**DOI:** 10.1101/2020.10.28.20221093

**Authors:** Shanshan Wan, Gaojing Qu, Hui Yu, Haoming Zhu, Guoxin Huang, Lei Chen, Meiling Zhang, Jiangtao Liu, Bin Pei

**Author notes:** Corresponding author **Correspondence:** Professor Jiangtao Liu, Department of Orthopedics, Xiangyang No.1 People’s Hospital, Hubei University of Medicine, Xiangyang 441000, Hubei Province, China. om. Professor Bin Pei, Center of Evidence-based Medicine, Xiangyang No.1 People’s Hospital, Hubei University of Medicine, Xiangyang 441000, Hubei Province, China.

## Abstract

**Objectives:** To study the features of creatine-kinase (CK) in COVID-19 patients with different ages, clinical types and outcomes and quantify the relationship between CK value and clinical type.

**Methods:** All laboratory confirmed COVID-19 patients hospitalized in Xiangyang No.1 People’s Hospital were included. Patients’ general information, clinical type, all CK values and outcome were collected.

**Results:** The peak median value of CK in cases aged ≥ 71 years old (appeared at T2) was higher than that in cases aged ≤ 70 years old. There was statistical difference between the two groups (*P*=0.001). Similarly, the peak in critical cases (appeared at T2) was higher than moderate and severe types, and significant difference were existed among moderate, severe, and critical types (*P*=0.000). Moreover, the peak value in death group (appeared at T2) was higher than those in survival group. Significant difference was also found between them (*P*=0.000). According to the optimal scale regression model, the CK value (*P*=0.000) and age (*P*=0.000) were associated with the clinical type.

**Conclusions:** Difference of the CK in different ages, clinical types, and outcomes were significant. The results of the optimal scale regression model are helpful to judge the clinical type of COVID-19 patients.

## 1. Introduction

Coronavirus disease-19(COVID-19), a virus induced pneumonia, has spread rapidly around the world[1-3]. Creatine-kinase (CK) is an important kinase directly related to intracellular energy transport, muscle contraction and ATP regeneration. It can reversibly catalyze the following reaction: ATP + creatine→ADP + phosphocreatine[4, 5], provide energy for muscle contraction and transport system[6]. CK was mainly found in the myocardium, skeletal muscle and brain, while little distributed in lung, gastrointestinal tract and thyroid. Sufficient CK can be released and terminally enhance the activity of these tissues under the pathological state[7]. Chen *et al*. indicated that CK abnormally increased in 13 (13.0%) out of 99 COVID-19 cases[8].

Two Meta analyses showed that the increase of CK value was significantly correlated to the severity of COVID-19 patients, and there was significant difference of CK value between severe and non-severe COVID-19 patients (*P*<0.01)[9, 10]. Zhou, Zheng *et al*. conducted a retrospective analysis of COVID-19 patients and indicated that CK value was significant higher in severe patients than those in mild patients. It was reported that the increase of CK could be served as a predictor of the severity of the disease[11, 12]. Chen Tao *et al*. described the clinical characteristics of 113 dead COVID-19 patients. According to their research, the median value of CK in the death group (113.0 U/L) was higher than that in the rehabilitation group (161.0 U/L)[13], but they did not conduct a statistical analysis of the differences. A univariate analysis showed that CK>185 U/L was associated with the death of COVID-19 patients[14].

At present, there is a lack of systematic research on the changes of CK value in COVID-19 patients with different ages, clinical types and outcomes, and the discrimination of clinical type with CK value still needs in-depth analysis. Therefore, we collected all the confirmed COVID-19 patients in Xiangyang No.1 People’s hospital, and analyzed the CK value since admission.

## 2. Methods

### 2.1 Study design and patients

This study was a bidirectional observational cohort study and the cohort was established on Feb 9, 2020. All suspected and confirmed cases of COVID-19 admitted to the Xiangyang No.1 People’s Hospital Affiliated Hospital of Hubei University of Medicine according to the Diagnosis and Treatment Protocol for Novel Coronavirus Pneumonia (1^st^-7^th^ editions) were included in this study[15]. The retrospective data were traced back to Jan 22, 2020 and the follow-up was carried out until Mar 28, 2020. Patients were classified into mild, moderate, severe, and critical type according to diagnosis and treatment protocol[15]. The classification results were cross-checked by two experts, and a third specialist involved if the inconsistency existed. The study was approved by the ethics review board of Xiangyang No.1 People’s Hospital (No. 2020GCP012) and registered at the Chinese Clinical Trial Registry as ChiCTR2000031088. Informed consent from patients has been exempted since this study does not involve patients’ personal privacy neither incur greater than the minimal risk.

### 2.2 Data collection

Data were extracted from the hospital information system by two groups and then cross-checked. Sex, age, CK value, disease onset date, outcome were collected.

### 2.3 Outcome measures

In this cohort, symptom onset was regarded as disease onset and the corresponding date was set as the day 1 to record data from symptom onset. Since the time from symptom onset to discharge was 26.87±9.19 days, we record the data from day 1 to day 30. The difference during day 5 to day 15 was most obvious, so we conduct statistical analysis on this period. We draw the CK median value/(5 days) trend chart with 5 days as the time unit. T1,T2…Tn was successively used to record per 5 days time unit, for example, T1 means the first 5 days after the onset of the disease. Mild type were excluded when we conducted the comparative and statistical analysis, because it only contained 4 cases.

### 2.4 Statistical analysis

All statistical analyses were performed through SPSS 20.0. Binary data were described by frequency and percentage. The normality of continuous data was checked. Mean and standard deviation were used to describe variables with normal distribution; otherwise, median (interquartile, IQR) was applied. Categorical data were described as frequency (%); the chi-square test was applied to assess significance between groups. Optimal scale regression model is used for regression analysis. All graphs were processed through GraphPad Prism 8.0 and Photoshop CC 14.2 software.

## 3. Results

This study included all of the suspected and laboratory-confirmed 542 patients till Feb 28, 2020. Among the 542 cases, the nucleic acid tests in 142 cases were positive. Excluding 2 infants, and 9 cases that transferred from other hospital whose data cannot be traced, 131 cases were included in the further study finally.

### 3.1 General information

Among the included 131 cases, there were 63 males and 68 females, and the average age was 50.1±17.1 years old. The average age was 48.2±16.2 and 73.1±9.5 years old in survival group and death group, respectively. The average time from onset to admission, from onset to discharge, from onset to death, and length of hospitalization were 4.5±3.1, 26.9±9.2, 18.4±9.8 and 22.4±8.7 days respectively. 121 cases were discharged from the hospital while 10 cases died.

### 3.2 CK test results

The 131 cases underwent 37 laboratory indicators contained 24052 tests in outpatient and hospitalization. This cohort included all of the CK results, totally 504 times of test, account for 2.1% of the all results of indicators. The normal range of CK value is 50.0-310.0 U/L. The change trend of CK median value showed a downward trend since the symptom onset (Supplement Figure A).

### 3.3 The features of CK in different ages

Cases were divided into 5 groups including ≤ 40, 41-50, 51-60, 61-70, and ≥ 71 years old group, while only the ≥ 71 years old group hold a obviously different distribution from other groups (Supplement Figure B). Moreover, significant difference of the CK median value was observed between the divided groups (*P*=0.000), while no statistical difference was found after excluding the ≥ 71 years old group (*P*=0.083) (Supplement Table). The age was correlated with CK value (*P*=0.000) with the coefficient as 0.235 in the five age groups (≤ 40, 41-50, 51-60, 61-70, ≥ 71 years old group); while no correlation was found between age and CK value after excluding the ≥ 71 years old group (*P*=0.344). Based on the above results, we divided the cases into ≤ 70, and ≥ 71 years old group.

In ≤ 70 years old group and ≥ 71 years old group, the CK median value was 50.9 U/L and 59.7 U/L, respectively, and the CK value abnormal percentage was 3.9% and 13.5%, respectively. The changes indicated that the CK median value in ≤ 70 years old group decreased during T1-T6, and the peak value (76.9 U/L) distributed at T1; in ≥71 years old group, the CK median value increased during T1-T2 and decreased after T2, and the peak value (205.7 U/L) distributed at T2. The CK median value all within normal range among the groups (Table 1, Fig. 1A).

**Table 1.** CK in COVID-19 patients with different age groups

| Period | ≤70 Years Old |  |  | ≥71 Years Old |  |  | P value |
| --- | --- | --- | --- | --- | --- | --- | --- |
|  | Median<br>(IQR) | N | Abnormal<br>Percentage<br>(%) | Median<br>(IQR) | N | Abnormal<br>Percentage<br>(%) | *P*=0.001 |
| Full Course | 50.9 (35.0-83.5) | 415 | 3.9% | 59.7 (35.4-203.9) | 89 | 13.5% | - |
| T1 | 76.9 (53.3-118.4) | 56 | 5.4% | 116.9 (84.9-312.1) | 11 | 27.3% | - |
| T2 | 64.6 (41.2-124.2) | 92 | 5.4% | 205.7 (89.2-710.6) | 21 | 33.3% | 0.000 |
| T3 | 48.4 (34.2-75.2) | 73 | 4.1% | 53.9 (38.9-75.5) | 17 | 5.9% | 0.291 |
| T4 | 45.9 (30.1-67.8) | 77 | 5.2% | 51.3 (28.2-67.6) | 19 | 0.0% | - |
| T5 | 43.6 (34.4-61.4) | 62 | 1.6% | 38.6 (32.3-48.7) | 14 | 0.0% | - |
| T6 | 37.7 (26.7-51.6) | 55 | 0.0% | 34.9 (24.1-45.7) | 7 | 14.3% | - |

N: total number of CK tests in corresponding period. *P*: P value between the two age groups during day 5 to day 15.

**Fig. 1.**
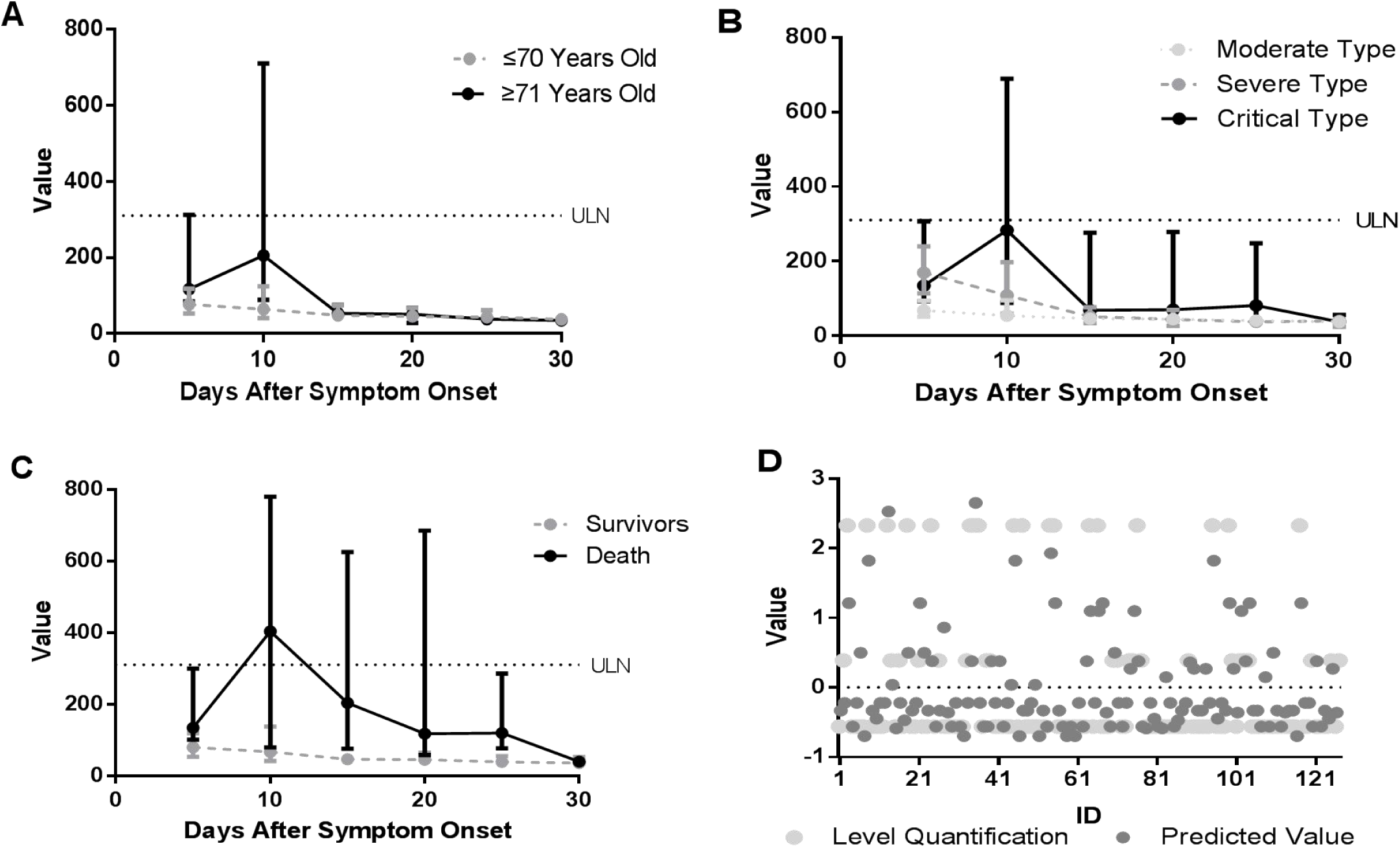
The change trend of CK median value in different ages (A), clinical types (B), outcomes (C), and comparison of quantitative scores of model output (D).

Statistical analysis of CK value from day 5 to day 15 showed statistical difference between the two groups (*P*=0.001), and the CK median value was higher in ≥ 71 years old group than those in ≤ 70 years old group.

### 3.4 The features of CK in different clinical types

In mild group, moderate group, severe group and critical group, the CK median value were 56.4 U/L, 46.2 U/L, 51.3 U/L and 79.50 U/L, respectively, and the abnormal percentage of CK value was 0.0%, 1.1%, 2.1% and 19.8%, respectively. The changes indicated that the CK median value of the moderate group and severe group decreased during T1-T6, and the peak value (67.4 U/L and 168.9 U/L, respectively) distributed at T1; in the critical group, the CK median value increased during T1-T2 and decreased after T2, and the peak value (283.0 U/L) distributed at T2. The CK median value all within normal range among the groups (Table 2, Fig. 1B).

**Table 2.** CK in COVID-19 patients with different clinical types

| Period | Moderate Type |  |  | Severe Type |  |  | Critical Type |  |  | P value |
| --- | --- | --- | --- | --- | --- | --- | --- | --- | --- | --- |
|  | Median<br>(IQR) | N | Abnormal<br>Percentage<br>(%) | Median<br>(IQR) | N | Abnormal<br>Percentage<br>(%) | Median<br>(IQR) | N | Abnormal<br>Percentage<br>(%) | *P=0.000* |
| Full Course | 46.2 (34.6-68.0) | 273 | 1.1% | 51.3 (32.6-114.3) | 95 | 2.1% | 79.5 (38.8-287.6) | 116 | 19.8% | - |
| T1 | 67.4 (50.9-93.4) | 41 | 2.4% | 168.9 (113.7-239.9) | 12 | 16.7% | 134.0 (92.1-307.0) | 10 | 30.0% | - |
| T2 | 53.9 (39.9-94.8) | 64 | 3.1% | 108.5 (59.1-197.0) | 21 | 0.0% | 283.0 (88.6-689.5) | 22 | 45.5% | *P1=0.043, P2=0.000, P3=0.003* |
| T3 | 45.6 (34.2-55.1) | 49 | 0.0% | 51.0 (33.4-76.6) | 17 | 0.0% | 68.3 (50.6-276.4) | 18 | 22.2% | *P2=0.003* |
| T4 | 44.6 (30.2-57.2) | 54 | 0.0% | 44.0 (26.5-68.9) | 20 | 0.0% | 69.5 (43.2-278.3) | 20 | 20.0% | - |
| T5 | 40.3 (29.3-50.7) | 37 | 0.0% | 37.1 (31.7-43.6) | 16 | 0.0% | 80.8 (38.7-247.7) | 22 | 4.5% | - |
| T6 | 37.2 (26.9-46.1) | 28 | 0.0% | 39.6 (23.6-50.9) | 9 | 0.0% | 37.2 (26.9-55.4) | 24 | 4.2% | - |

N: total number of CK tests in corresponding period. *P*: P value among the three clinical types during day 5 to day 15. *P1*: P value between moderate type and severe type in corresponding period. *P2*: P value between moderate type and critical type in corresponding period. *P3*: P value between severe type and critical type in corresponding period.

**Table 3.** CK in COVID-19 patients with different outcomes

| Period | Survival Group |  |  | Death Group |  |  | P value |
| --- | --- | --- | --- | --- | --- | --- | --- |
|  | Median<br>(IQR) | N | Abnormal<br>Percentage<br>(%) | Median<br>(IQR) | N | Abnormal<br>Percentage<br>(%) | *P*=0.000 |
| Full Course | 48.9 (34.0-80.4) | 453 | 2.6% | 138.7 (59.7-470.0) | 49 | 32.7% | - |
| T1 | 79.4 (53.3-119.3) | 60 | 6.7% | 134.0 (101.1-299.4) | 6 | 33.3% | - |
| T2 | 67.3 (41.5-137.8) | 100 | 5.0% | 403.4 (79.8-779.4) | 12 | 58.3% | 0.001 |
| T3 | 46.2 (34.2-61.1) | 81 | 1.2% | 203.9 (75.5-625.3) | 9 | 33.3% | 0.000 |
| T4 | 45.0 (29.9-64.8) | 87 | 1.1% | 117.5 (58.6-685.2) | 9 | 33.3% | - |
| T5 | 39.1 (31.5-55.3) | 69 | 1.4% | 119.4 (77.0-285.7) | 7 | 0.0% | - |
| T6 | 35.9 (26.2-52.2) | 56 | 0.0% | 39.8 (36.2-48.9) | 6 | 16.7% | - |

N: total number of CK tests in corresponding period. *P*: P value between the two outcome groups during day 5 to day 15.

Statistical analysis of CK value from day 5 to day 15 showed statistical difference among the three clinical type groups (*P*=0.000). The CK median value in severe type was significantly higher than those in moderate type (*P*=0.006). The CK median value in critical type was significantly higher than those in moderate type (*P*= 0.000) and severe type (*P*=0.004).

The maximum CK in the first 15 days after symptom onset was collected to build the categorical regression based on optimal scale. For the reason that the mild type only contained 4 cases, we excluded this type. The model output showed that age (*P*=0.000) and CK (*P*=0.000) had significant correlation while sex (*P*=0.253) did not have significant correlation with clinical type. The quantification of CK categories showed CK ≤ 310.33 U/L was related to moderate type. CK ranged 360.37-883.44 U/L was related to severe type, and CK > 883.44 U/L was related to critical type. The quantification of age showed ≤ 61 years old was related to moderate type, 62-70 years old was related to the severe type, and 71-90 years old was related to critical type (Table 4). The model expression was Q_level= 0.311* Q_CK+0.525* Q_age+ 0.059* Q_sex (Q_level, Q_CK, Q_age and Q_sex represent the scale quantization score of clinical type, CK value, age and sex, respectively). The comparison between severity quantification of different clinical type output by this model and the actual severity quantification of the clinical type classified by the respiratory physicians was shown in Fig.1D. The accuracy rate of this output on three types (moderate, severe, critical) was 73.6%. The value of the model below 1 is divided into moderate type and severe type, and over 1 is divided into critical type because only the critical type hold a obviously different distribution from other clinical types (Fig. 1B). The accuracy of distinguishing the two types is 89.6%.

**Table 4.** Variable categories and quantification score in optimal scale regression model

| Variable | Classification | Frequency | Quantification |
| --- | --- | --- | --- |
| Clinical Type | Moderate Type | 86 | -0.571 |
|  | Severe Type | 21 | 0.382 |
|  | Critical Type | 18 | 2.284 |
| CK | 4.05-61.08 | 43 | -0.705 |
|  | 63.01-310.33 | 70 | -0.043 |
|  | 360.37-457.70 | 6 | 2.229 |
|  | 710.57-883.44 | 3 | 2.229 |
|  | 1127.92-4371.26 | 3 | 4.427 |
| Sex | Male | 60 | 1.041 |
|  | Female | 65 | -0.961 |
| Age | 15-29 | 16 | -0.778 |
|  | 30-39 | 20 | -0.536 |
|  | 40-47 | 18 | -0.536 |
|  | 48-54 | 20 | -0.536 |
|  | 55-61 | 15 | -0.536 |
|  | 62-70 | 18 | 0.683 |
|  | 71-90 | 18 | 2.184 |

### 3.5 The features of CK in different outcomes

In survival group and death group, the CK median value was 48.9 U/L and 138.7 U/L, respectively, and the CK value abnormal percentage was 2.7% and 32.7%, respectively. The changes indicated that the CK median value in survival group decreased during T1-T6, and the peak value (79.4 U/L) distributed at T1; while in death group, the CK median value increased during T1-T2 and decreased after T2, and the peak value (403.4 U/L) distributed at T2, the abnormal time interval was T2 (Table 3, Fig. 1C).

Statistical analysis of CK value from day 5 to day 15 showed statistical difference between the two outcome groups (*P*=0.000), and the CK value in death group was higher than those in survival group. According to the spearman correlation analysis, the correlation was found between outcome and CK value (*P*=0.000) with the coefficient as 0.341 in the two outcome groups.

## 4. Discussion

The CK value in cases aged ≥ 71 years old group increased during day 5 to day 15, which was significantly different from other age groups. The correlation analysis suggested a correlation between the ≥ 71 years old group and the CK value, indicating that among the five age groups, cases aged ≥ 71 years old had the most severe cell injury and induced the increased CK value. CK is mainly distributed in high energy metabolic organs such as heart and brain, indicating that important organs are more likely to be damaged in cases aged ≥ 71 years old. This is consistent with the result of our regression analysis that 71-90 years old was related to critical type (Table 4). It is suggested that we should pay close attention to the changes of CK value in cases aged ≥ 71 years old, and this is consistent with the multivariate regression analysis conducted by Zhou *et al*, which showed that the death of COVID-19 patients usually associated with the advanced age[14]. But the reason that the important hypermetabolic organs in cases aged ≥ 71 years old are prone to serious damaged worth further study.

There were significant differences among different clinical types, and the more serious the type, the higher the peak value of CK, and the greater the proportion of CK over ULN. At T2, significant differences were found in the CK value among the three clinical types, suggesting that T2 may be the period for judging the clinical type of COVID-19 patients, CK during T2 period should be closely monitored, and CK > 197.0 U/L (upper quartile for severe type) may develop into critical type. Therefore, we recommend that clinicians should closely monitor the CK value in the early stage of COVID-19 patients for early warning and intervention. In order to quantify the relationship between CK value and clinical type, regression analysis was conducted based on CK, age, and sex. Since the time from onset to severe type and critical type is 8.15 ± 5.29 days and 11.83 ± 5.19 days respectively, the maximum values of CK in the first 15 days were selected in this model. The results show that the CK value can be used to distinguish clinical types, which can help us to grasp the opportunity of treatment better and prevent patients from developing into critical type. However, the prediction ability of this model need to be verified by other data.

According to the difference between different outcomes, we found that the death group had higher peak value of CK and the greater proportion of CK exceeding ULN. Statistical analysis showed that there was statistical difference between the two outcome groups. Spearman correlation analysis showed that outcome was correlated with CK value. At T2 and T3, the CK value of the death group was significantly higher than that of survival group (*P*=0.001, *P*=0.000 respectively), suggesting that T2 might be the first period to judge the outcome of the disease. And CK > 137.8 U/L (upper quartile of survival group) has a risk of death. This is of reference value for us to judge the prognosis, especially in the early stage.

To sum up, patients in different ages, clinical types and outcomes, especially those ≥ 71 years old, critical and dead patients, the CK median value reached the peak at the day 10, which appeared early and concentrated, and then decreased rapidly. It is suggested that the day 10 may be a good index for early clinical classification, and the most critical time point in the progression of the disease. CK exists in a large number of cells which rapidly regenerated by ATP, such as heart muscle, skeletal muscle and brain tissue, and these tissues are important to maintain the normality of human signs. Once the tissues damaged, the permeability of cell membrane increased, which makes CK continuously released into the blood, and induced the increased CK value finally[16]. Therefore, early monitoring of CK value has important reference value for us to judge the severity of the disease and the outcome of the disease.

## 5. Limitation

The current study has several limitations. First, the cases included in our cohort were enrolled from single hospital, the evidence value was limited. Second, due to the limited sample size, the data could not be analyzed and described daily, and some information has been lost. Third, multicenter and large sample studies were needed to verify the optimal scale regression model based on clinical type.

## 6. Conclusion

The results showed that the more serious the clinical type, the higher the peak value of CK. The peak value of CK appeared at day 10 in the ≥ 71 years old group, critical type and death group, which appeared early and concentrated, indicated that early monitoring of CK value is helpful to better evaluate the disease and is of great significance to the graded treatment of patients, the day 10 after onset (4-5 days after hospitalization) may be the most critical time point. The results of the optimal scale regression model are helpful to judge the clinical type of COVID-19 patients. Spearman correlation analysis showed that outcome was correlated with CK value. From the characteristics of changes in CK value, and our model, we can make up for the deficiency of CK in the early diagnosis, severity assessment and prognosis judgment of COVID-19 patients.

## Data Availability

Anyone who wishes to obtain the original data of this study with reasonable purposes can contact the correspondent author via email.

## 7. Declaration

### 7.1 Ethics approval and consent to participate

The study was approved by the ethics review board of Xiangyang No.1 People’s Hospital (No. 2020GCP012) and registered at the Chinese Clinical Trial Registry as ChiCTR2000031088. Informed consent from patients has been exempted since this study does not involve patients’ personal privacy neither incur greater than the minimal risk.

### 7.2 Funding this research

This research did not receive any specific grant from funding agencies in the public, commercial, or not-for-profit sectors.

### 7.3 Conflict of interests

The authors have no competing interest to declare.

## 7.4 Acknowledgement

We want to thank all medical workers for their great contribution and sacrifice during this COVID-19 pandemic.

---

**Supplement Table.**
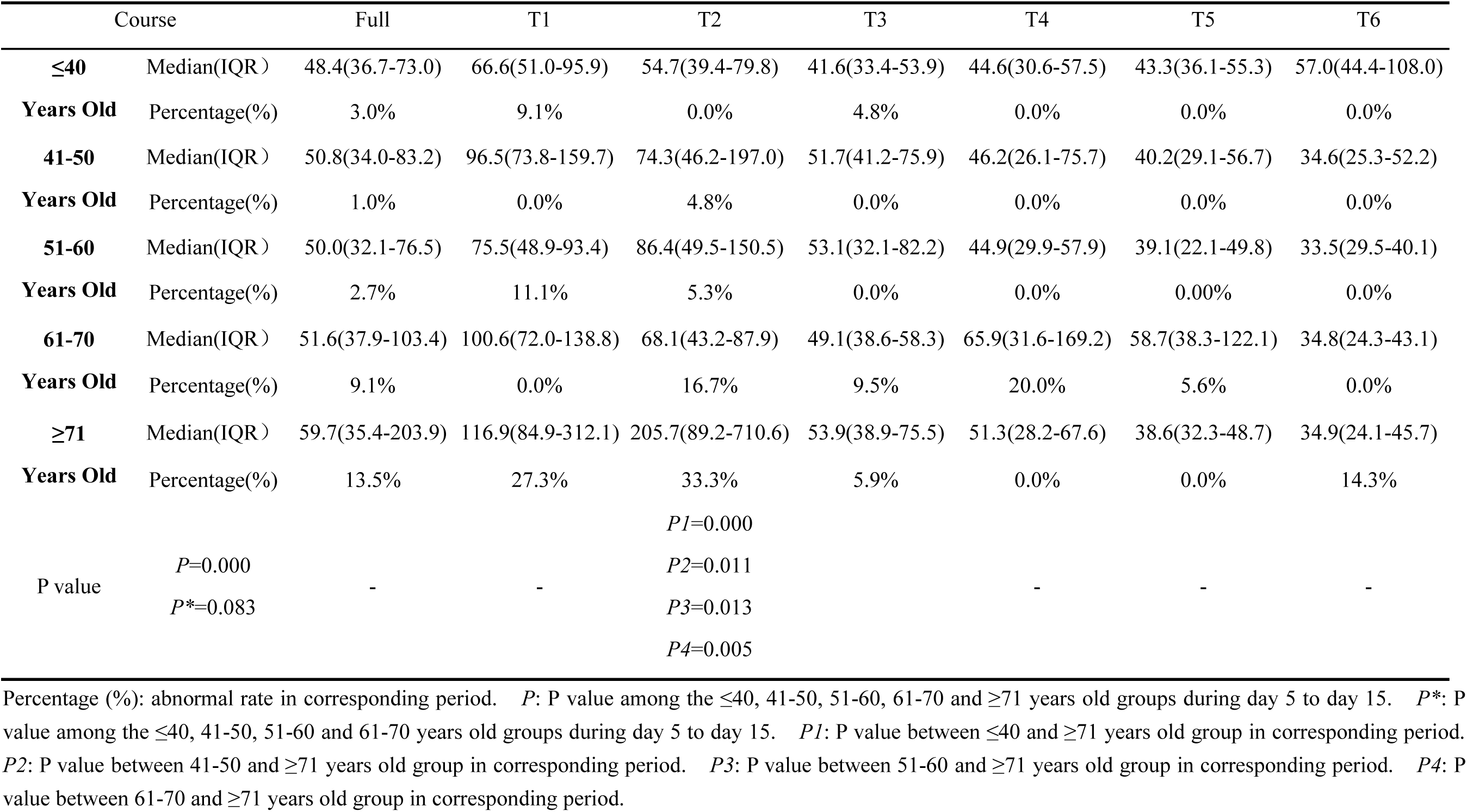
CK in COVID-19 patients with five age groups

**Supplement Figure.**
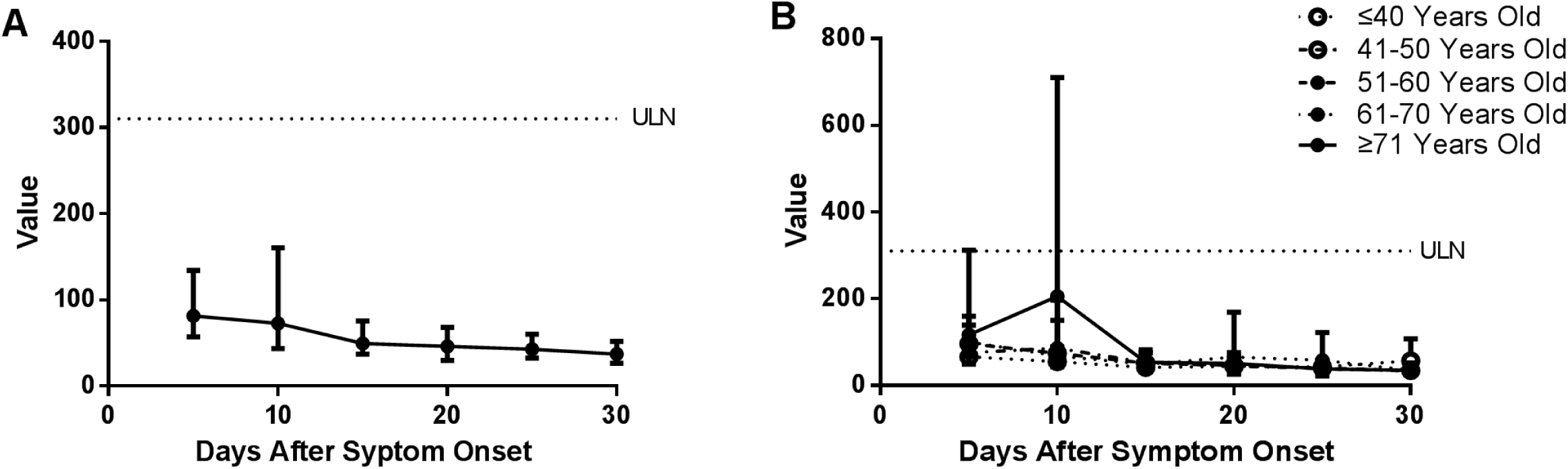
The change trend of CK median value in 30 days (A) and in five age groups (B)

## Notes

### Competing Interest Statement

The authors have declared no competing interest.

### Clinical Trial

ChiCTR2000031088

### Funding Statement

No external funding was received in this study.

### Author Declarations

This study was unanimously approved by Ethics Committee of Xiangyang NO.1 people's hospital on February 12, 2020.

